# Efficacy and Safety of Transcranial Temporal Interference Stimulation for Improving Negative Symptoms and Cognition in Schizophrenia: A Pilot Study

**DOI:** 10.1101/2025.03.31.25324806

**Authors:** Shuzhe Wang, Nannan Liu, Junhao Chen, Linxuan Wang, Jingxuan Liu, Lijun Wang, Mingjin Wang, Shen Li, Jie Li

**Author notes:** These authors contributed equally to this work and should be considered co-first authors (Shuzhe Wang, Nannan Liu). Corresponding authors: Jie Li, M.D., PhD.; Institute of Mental Health, Tianjin Anding Hospital, Mental Health Center of Tianjin Medical University, No. 13, Liulin Road, Hexi District, Tianjin 300222, China, Shen Li, M.D., PhD.; Institute of Mental Health, Tianjin Anding Hospital, Mental Health Center of Tianjin University, No. 13, Liulin Road, Hexi District, Tianjin 300222, China.

## Abstract

**Importance:** Schizophrenia (SCZ) is a severe psychiatric disorder with limited treatment options for negative symptoms and cognitive impairment. Temporal interference stimulation (TIS) is a non-invasive neuromodulation technique that enables targeted deep brain modulation. However, its therapeutic effects on SCZ remain unexplored.

**Objective:** To evaluate the safety, efficacy, and tolerability of TIS targeting the right nucleus accumbens (NAc) in improving negative symptoms and cognitive function in patients with SCZ.

**Design:** An open-label, single-arm exploratory trial conducted from July 20 to October 30, 2024.

**Setting:** Single-center study conducted at a tertiary psychiatric referral hospital (Tianjin Anding Hospital, China).

**Participants:** 8 SCZ inpatients (mean age 48.3 ± 12.1 years; 6 males) with persistent negative symptoms (PANSS negative subscale ≥20) were enrolled.

**Intervention:** Five consecutive daily sessions of 20-minute individualized MRI-guided 130 Hz TIS targeting the right NAc.

**Main Outcomes and Measures:** Primary outcomes: changes in PANSS negative subscale and SANS scores from baseline to two-week follow-up. Secondary outcomes: changes in MATRICS Consensus Cognitive Battery (MCCB) scores across assessment phases. Assessments occurred at baseline, post-intervention, and two-week follow-up.

**Results:** All 8 participants completed treatment. PANSS total scores decreased by 8.1% (baseline: 64.5 ± 14.8 vs follow-up: 59.3 ± 15.7; p_adj_ = 0.007), driven by reductions in negative (baseline: 25.4 ± 6.7 vs follow-up: 23.0 ± 6.7; p_adj_ = 0.045) and general subscales. SANS scores declined by 12.5% (baseline: 57.8 ± 9.4 vs follow-up: 50.6 ± 12.9; p_adj_ = 0.045). MCCB total scores improved by 41.8% (baseline: 23.7 ± 10.4 vs follow-up: 33.6 ± 9.7, p_adj_ < 0.001), with significant gains in visual and verbal learning (p_adj_ ≤ 0.02). Adverse events included electrode contact site pressure sensation (n = 4), transient current sensations (n = 4) and fatigue (n = 1). Tolerability was good.

**Conclusion:** This pilot study suggests that TIS is a safe and feasible noninvasive deep brain stimulation technology that may alleviate negative symptoms and improve cognitive function in SCZ. Larger randomized controlled trials with extended follow-up periods are warranted to further validate these preliminary results.

**Trial Registration:** Chinese Clinical Trial Registry; ChiCTR2400094200.

## Introduction

Schizophrenia (SCZ) is a severe and progressive psychiatric disorder characterized by positive, negative, and cognitive symptoms^1^. Among these, negative symptoms and cognitive impairment are considered core features of SCZ, contributing significantly to functional disability and poor long-term outcomes^2^. Despite being the mainstay of treatment, current antipsychotic medications show limited efficacy in alleviating these core symptoms^3^.

Recent advances in neuromodulation have indicated that deep brain stimulation (DBS) has the potential to function as a potential therapeutic intervention for refractory SCZ^4^. Clinical trials targeting the nucleus accumbens (NAc), subgenual anterior cingulate cortex (sgACC), and habenula have reported significant reductions in negative symptoms and improvements in affective processing in patients with SCZ^2,5^. However, the clinical translation of DBS is hindered by its invasive nature, which entails risks of surgical complications (e.g., hemorrhage, infection), hardware-related adverse events (e.g., lead displacement), and long-term device maintenance burdens ^6,7^. These limitations underscore the need for non-invasive alternatives capable of achieving comparable therapeutic precision.

To address these challenges, Grossman et al. proposed temporal interference stimulation (TIS), a non-invasive neuromodulation technique that enables precise targeting of deep brain structures^8^. TIS utilizes two pairs of scalp electrodes to deliver high-frequency alternating currents (e.g., 2,000 Hz and 2,010 Hz). The interference of these fields generates a low-frequency amplitude-modulated envelope wave (e.g.,10 Hz) within the brain, which can be spatially focused on deep brain regions such as the hippocampus and striatum^9,10^. Crucially, the high-frequency carrier waves bypass neuronal activation thresholds, thereby avoiding unintended stimulation of superficial cortical areas^8^. Compared to conventional transcranial electrical stimulation, TIS achieves deeper penetration while maintaining comparable spatial specificity. Additionally, unlike DBS, TIS eliminates surgical risks associated with invasive procedures. To date, TIS has shown promise in modulating neural activity in both healthy populations^11,12^ and patients with bipolar disorder^13^, Parkinson’s ^7^and Alzheimer’s disease^14^. However, its therapeutic potential in SCZ remains unexplored.

In the present study, we focused on the NAc as a therapeutic target in SCZ. The rationale for targeting the NAc is twofold. Firstly, neurobiological evidence indicates that SCZ patients exhibit pathological NAc hyperactivity, characterised by elevated dopamine levels^15^. Theoretically, this hyperdopaminergic state is linked to core symptom domains, including aberrant salience attribution (positive symptoms)^16^, blunted reward processing (negative symptoms)^17^, and disrupted prefrontal cortical connectivity (cognitive impairments)^18^. Thus, modulating NAc hyperactivity may concurrently alleviate this tripartite symptom spectrum. Secondly, previous research conducted using the DBS technique has demonstrated that targeting the NAc has produced symptomatic improvements in patients diagnosed with SCZ, which further underscores its therapeutic potential^5^.

In this pilot study, we aim to investigate the safety, efficacy and tolerability of TIS on right NAc in SCZ patients with both severe negative symptoms and cognitive dysfunction.

## Materials and methods

### Study design

This study was designed as an open-label, exploratory trial conducted at the Brain Assessment & Intervention Laboratory, Tianjin Anding Hospital. The study protocol (Fig. 1) received approval from the Ethics Committee of Tianjin Anding Hospital. All procedures conformed to the ethical principles established in the Declaration of Helsinki for human medical research. The trial was registered at the Chinese Clinical Trial Registry (www.chictr.org.cn, ChiCTR2400094200).

**Fig 1.**
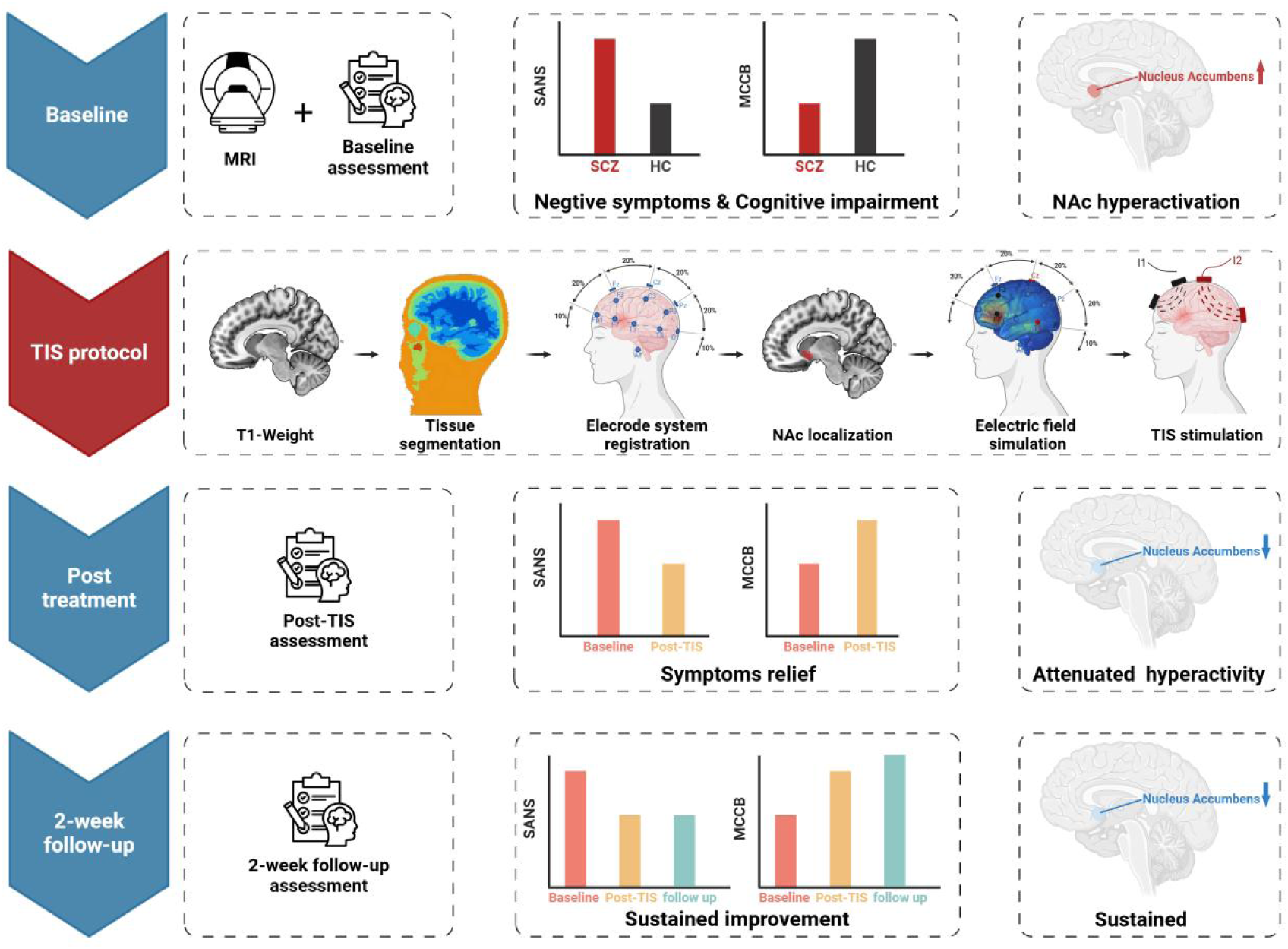
Graph Abstract.

### Participants

Participants were recruited from the inpatient units of Tianjin Anding Hospital between July 20, 2024, and October 30, 2024. The inclusion criteria were as follows: individuals aged 18–65 years (inclusive); a confirmed diagnosis of schizophrenia (either first episode or relapse) according to the Diagnostic and Statistical Manual of Mental Disorders, Fifth Edition (DSM-5); a Negative Symptom Subscale score ≥ 20 on the Positive and Negative Syndrome Scale (PANSS)^19^; a stable antipsychotic regimen for at least 30 days prior to enrollment; and adequate listening, speaking, reading, and writing abilities to complete study assessments. Exclusion criteria included: any co-existing psychiatric disorders, neurological conditions, or substance use disorders that could influence study outcomes; a history of seizures or epileptic convulsions; the presence of metallic foreign bodies in the skull or metallic implants in the heart; a history of organic brain disorders, severe head trauma, or brain surgery; receipt of electroconvulsive therapy or other physical treatments (e.g., transcranial magnetic or electrical stimulation) within 30 days before enrollment; any changes in antipsychotic medication within the last 30 days prior to enrollment; a score of ≥ 3 on the suicide item of the 17-item Hamilton Depression Scale; pregnancy, breastfeeding, or plans to conceive during the study period; and concurrent participation in other interventional clinical trials. All participants provided written informed consent before joining the study.

### Modeling

High-resolution three-dimensional structural MRI images were acquired using a 3.0 Tesla Siemens Prisma scanner (Erlangen, Germany) at Tianjin Anding Hospital. A T1-weighted MPRAGE sequence was employed with the following parameters: repetition time (TR) = 2000 ms; echo time (TE) = 2.32 ms; flip angle = 8^°^; field of view (FOV) = 256 × 256 mm²; voxel size = 1 × 1 × 1 mm³; and 208 sagittal slices.

T1-weighted MRI images were segmented using SPM12 (http://www.fil.ion.ucl.ac.uk/spm/) into six tissue types: scalp (0.465 S/m), skull (0.01 S/m), cerebrospinal fluid (1.65 S/m), white matter (0.126 S/m), gray matter (0.276 S/m), and cavity (2.5 × 10⁻¹⁴ S/m). The 10-10 EEG electrode system was then registered onto each participant’s scalp to establish a standardized electrode placement framework.

Next, tetrahedral finite element meshes of the head were generated using the Computational Geometry Algorithms Library (CGAL)^20^. The finite element solver GetDP^21^ was used to compute the electric field intensities for each electrode pair. By applying the TIS electric field coupling principle, we calculated and analyzed the electric field distribution for each participant.

To localize the right NAc, the AAL3 template^22^ was registered to each participant’s MRI images. An exhaustive search algorithm was then implemented to identify the optimal electrode configurations that would maximize electric field intensity at the right NAc. These individualized electrode configurations were subsequently utilized in the stimulation protocol.

### TIS stimulation paradigm

TIS was delivered using the NervioX-1000 device (SuZhou NeuroDome Clinical Technology Co., Ltd.), which includes a stimulator console, electrode adapter, integrated stimulation algorithms, and a cap with cables and electrodes. The device met international safety and electromagnetic compatibility (EMC) standards, verified by EPINTEK Suzhou.

Electrodes were placed according to the 10-10 EEG system and secured with Ten20 conductive paste (Weaver and Company, Aurora, CO, USA). Each electrode pair delivered continuous high-frequency sinusoidal currents at 2000 Hz and 2130 Hz, respectively, producing a beat frequency of 130 Hz, as guided by previous DBS research^5^.

Each stimulation session lasted for 20 minutes, with an additional 10-second ramp-up and ramp-down period at the start and end of each session. The treatment protocol consisted of five consecutive daily sessions (one session per day). The initial treatment session for each patient was documented on video.

### Assessment

Clinical assessments were conducted at baseline, post-intervention, and during the two-week follow-up. Psychotic symptoms were assessed using the PANSS. Negative symptoms were systematically measured with the Scale for the Assessment of Negative Symptoms (SANS)^23^. Cognitive function was evaluated using the MATRICS Consensus Cognitive Battery (MCCB)^24^. A 10-point visual analogue scale (VAS), ranging from 0 ("no sensation") to 10 ("maximum tolerance") similar to pain assessment scales, was employed to evaluate TIS tolerability. All clinical assessments were administered by trained master’s-level psychiatrists. All adverse events during the treatment were recorded.

### Statistical analysis

A repeated measures analysis of variance (RM-ANOVA) was employed to assess clinical scale scores across three time points: baseline, post-intervention, and the two-week follow-up. Where Mauchly’s test indicated violations of sphericity, the Greenhouse-Geisser adjustment was implemented. The Benjamini-Hochberg procedure was applied for multiple comparison correction. Statistical significance was defined as p < 0.05. Statistical analyses were performed using SPSS 22.0 (IBM SPSS Statistics, Chicago, IL).

## Results

### Participants

A total of eight patients (six males and two females) were enrolled, with a mean age of 48.3 ± 12.1 years, education duration of 12.7 ± 2.3 years, illness duration of 23.4 ± 11.8 years, and current episode duration of 5.6 ± 3.7 years. Detailed demographic and medication information are provided in (Table 1).

**Table 1.** Sociodemographic characteristics and medication history.

| Patient | Age<br>range | Sex<br>(M/F) | Education<br>(Years) | Duration of<br>Illness<br>(Years) | Duration Current<br>Episode<br>(Years) | Medications<br>(mg/day) <sup>a</sup> |
| --- | --- | --- | --- | --- | --- | --- |
| 1 | 41-45 | M | 12 | 20 | 3 | Olanzapine 7.5<br>Bupropion 16<br>Sertraline 100<br>Zolpidem 5 |
| 2 | 36-40 | M | 16 | 10 | 7 | Aripiprazole 15<br>Olanzapine 5 |
| 3 | 26-30 | F | 9 | 12 | 1 | Aripiprazole 10<br>Olanzapine 20<br>Trihexyphenidyl 6 |
| 4 | 61-65 | M | 12 | 37 | 12 | Clozapine 250 |
| 5 | 61-65 | M | 12 | 12 | 9 | Olanzapine 2.5<br>Alprazolam 0.4 |
| 6 | 56-60 | M | 16 | 36 | 5 | Risperidone 6<br>Trihexyphenidyl 6<br>Zopiclone 7.5<br>Lorazepam 1 |
| 7 | 41-45 | M | 13 | 23 | 2 | Lurasidone 40<br>Clozapine 350<br>Lorazepam 1.5<br>Sodium Valproate 1000 |
| 8 | 51-55 | F | 12 | 37 | 6 | Olanzapine 12.5 |

Individualized electrode configurations were optimized through electric field modeling. Two electrode pairs (I1 and I2) generated electric field intensities within the target region for each patient, ranging from 0.68 to 1.54 V/m. The stimulation currents maintained between 2.3 to 4.0 mA (AC peak-to-baseline). Three-dimensional electric field distributions and stimulation parameters are presented in (Supplemental Fig. 1 and Supplemental Table 1).

### Safety and tolerability

During the course of treatment, four patients reported experiencing “electrode contact site pressure sensation”, while four others reported “transient current sensations”. These sensations were transient and did not require intervention. The tolerability scores of TIS were 7.6 ± 1.0 (assessed via the VAS), with scores in the 6-8 range corresponding to perceptible yet tolerable stimulation intensity. All participants completed five treatment sessions, with only one case (patient 1) of transient fatigue reported post-treatment. No other adverse events were observed across the cohort. The first session for each patient was video-recorded.

### Clinical outcomes

The PANSS total scores demonstrated a decrease from 64.5 ± 14.8 at baseline to 59.5 ± 14.9 post-treatment (7.8%), and further decline to 59.3 ± 15.7 at follow-up (8.1%). Time-course analysis revealed significant main effects (F = 7.8, p = 0.02), with notable differences observed between baseline and post-treatment (p_adj_ = 0.007) as well as baseline and follow-up (p_adj_ = 0.007) (Table 2 and Fig. 2A). For the subscales, both the General and Negative Subscales exhibited significant main effects over time (General Subscale: F = 5.3, p = 0.03; Negative Subscale: F = 3.9, p = 0.05). Specifically, scores on the General Subscale declined from 28.1 ± 6.6 at baseline to 26.0 ± 5.8 post-treatment (p_adj_ = 0.04), while the Negative Subscale showed a reduction from 25.4 ± 6.7 at baseline to 23.0 ± 6.7 at follow-up (p_adj_ = 0.045). No significant changes were observed in the Positive Subscale over time (p > 0.05) (Fig. 2B-D).

**Fig 2.**
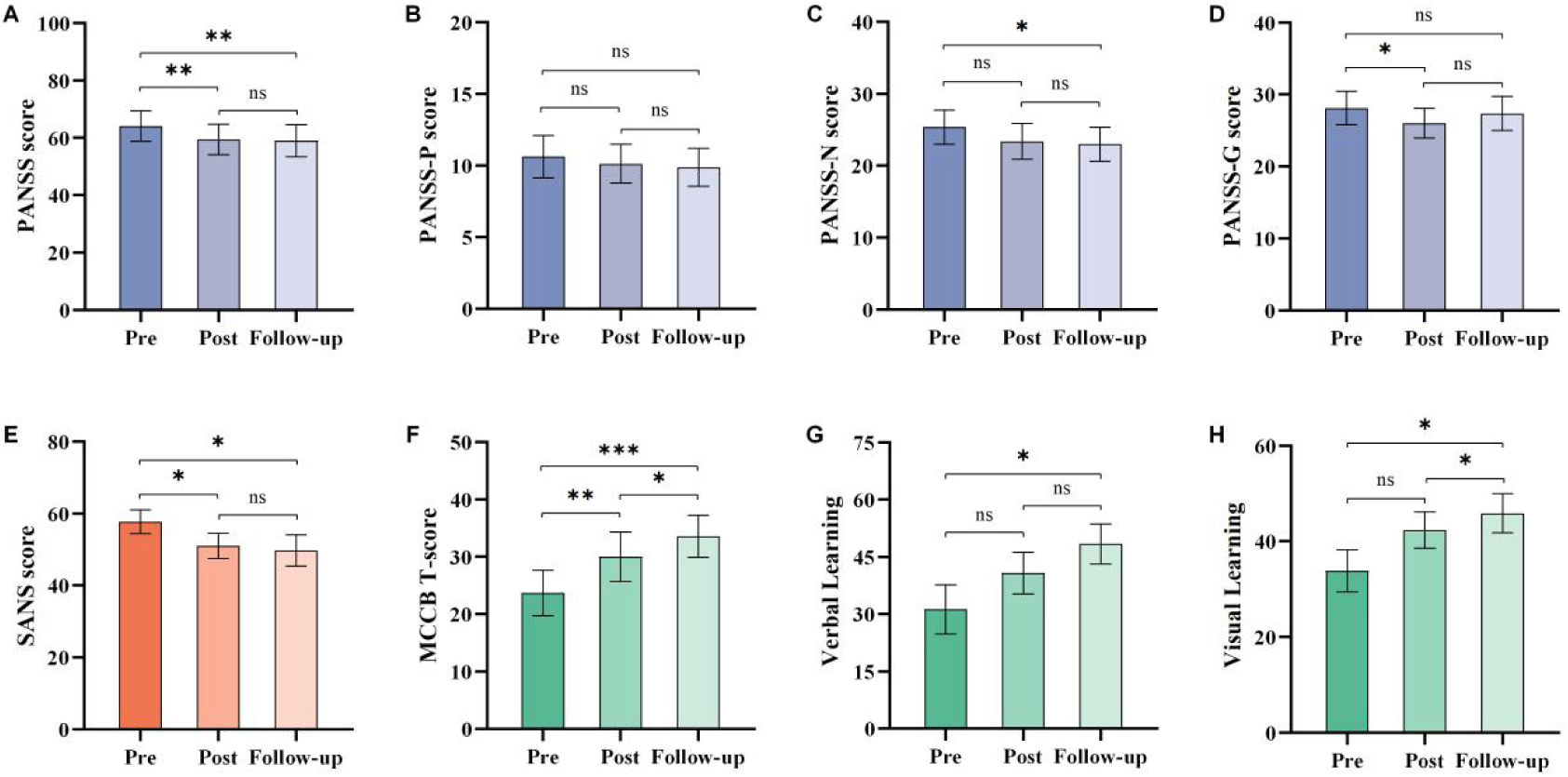
Effects of temporal interference stimulation on negative symptoms and cognitive dysfunction. (A-D) PANSS total and subscales scores. (E) Total score of SANS. (F-H) Total score of MCCB and subdomains of Visual Learning and Verbal Learning. PANSS: Positive and Negative Syndrome Scale; SANS: Scale the Assessment of Negative Symptoms; MCCB: MATRICS Consensus Cognitive Battery. *Significant at p_adj_ < 0.05. **Significant at p_adj_ < 0.01. ***Significant at p_adj_ < 0.001.

**Table 2.**
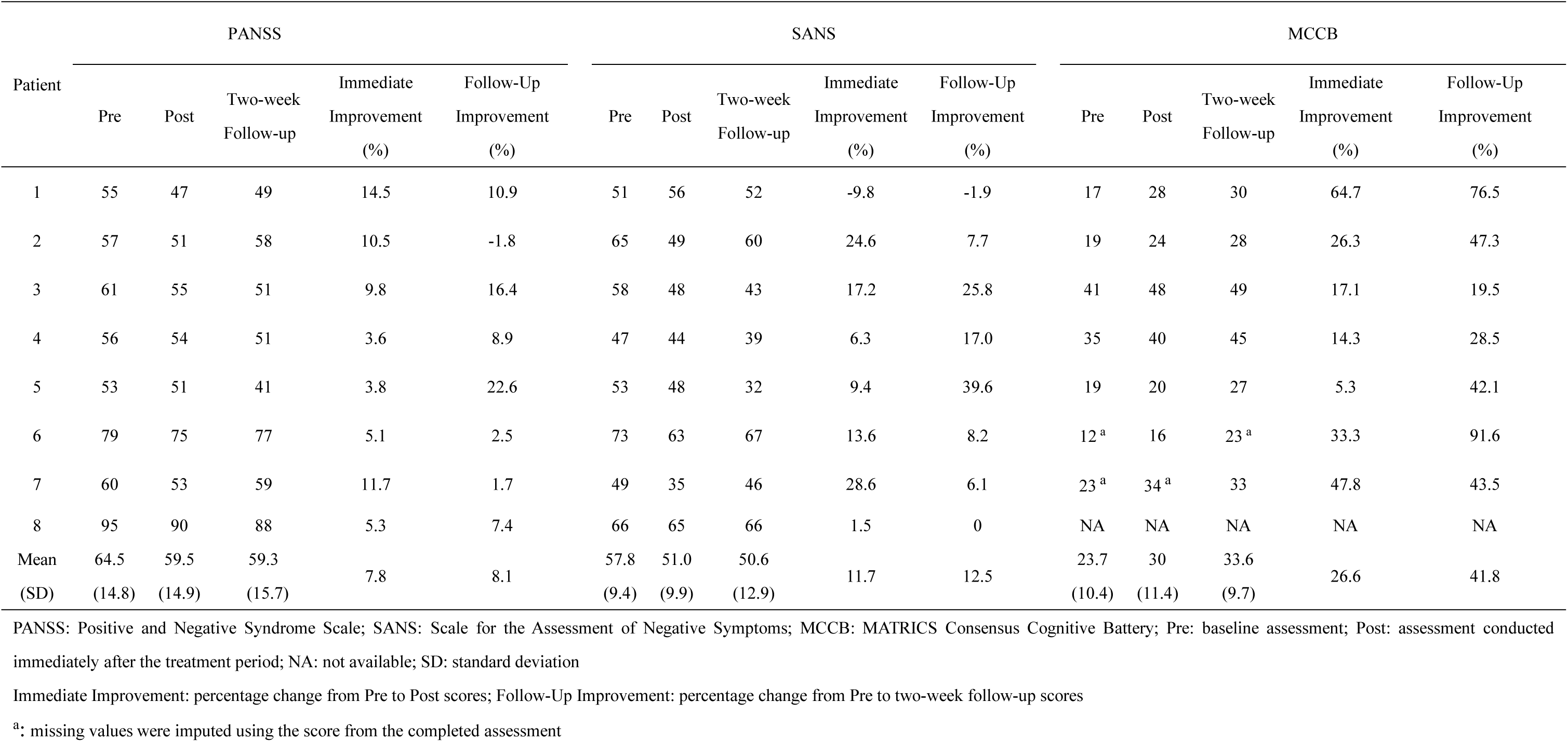
Outcome measures before, after TIS stimulation, and at two-week follow-up.

SANS total scores decreased from 57.8 ± 9.4 at baseline to 51.0 ± 9.9 post-treatment (11.7%), followed by a further decrease to 50.6 ± 12.9 at follow-up (12.5%). Time-course analysis revealed a significant main effect of time (F = 5.1, p = 0.02), with post-hoc comparisons indicating statistical significant reductions from baseline to both post-treatment (p_adj_ = 0.045) and follow-up (p_adj_ = 0.045) (Table 2 and Fig. 2E). Subscale analysis demonstrated a downward trend in scores, though this did not reach statistical significance (Supplemental Fig. 2).

### Cognitive outcomes

In the present study, five patients completed all MCCB assessments. Two participants had complete data at one timepoint but missing social cognition scores later (imputed from existing data), and one was excluded due to incomplete data.

The MCCB total scores demonstrated an improvement from 23.7 ± 10.4 at baseline to 30.0 ± 11.4 post-treatment (26.6%), and further improvement to 33.6 ± 9.7 at follow-up (41.8%). Time-course analysis revealed significant main effects (F = 40.2, p < 0.001), with notable differences observed between baseline and both post-treatment (p_adj_ = 0.006) and follow-up (p_adj_ < 0.001), as well as between post-treatment and follow-up (p_adj_ = 0.02) (Table 2 and Fig. 2F). Regarding the subdomains, both Visual Learning and Verbal Learning exhibited significant main effects over time (Visual Learning: F = 7.2, p = 0.03; Verbal Learning: F = 7.6, p = 0.01). Specifically, Visual Learning scores improved from 33.9 ± 12.5 at baseline to 45.9 ± 11.7 at follow-up (p_adj_ = 0.02), while Verbal Learning scores increased from 31.3 ± 18.1 at baseline to 40.8 ± 15.4 post-treatment and further to 48.4 ± 14.8 at follow-up (p_adj_ = 0.02) (Fig. 2G-H).

## Discussion

To the best of our knowledge, this is the first study to evaluate the efficacy and safety of TIS in patients with SCZ. Despite the limited sample size, our preliminary findings demonstrated that 130Hz TIS targeting right NAc could improve both negative symptoms and cognitive function in SCZ patients. These findings indicate that TIS is a promising non-invasive therapeutic strategy for patients with SCZ.

Our results demonstrated an improvement in negative symptoms by targeting the NAc, as evidenced by significant reductions in SANS and PANSS negative subscale scores. This finding aligns with previous DBS studies, which have shown that stimulation of the NAc can alleviate negative symptoms in patients with SCZ^5^, further supporting the therapeutic potential of modulating this brain region. However, it should be note that although a downward trend was observed in the SANS subdimensions, these changes did not reach statistical significance—likely due to the small sample size and short treatment duration. Additionally, no significant improvement in positive symptoms was observed during the study, which may be attributed to the fact that enrolled patients had already achieved sufficient relief from these symptoms prior to enrollment. Moving forward, neuroimaging studies are needed to clarify TIS-induced changes in NAc activity and to better understand the mechanisms underlying these therapeutic effects.

The present study demonstrated a significant improvement in cognitive function following TIS, as reflected by the increase in MCCB total scores. This finding is consistent with mounting evidence that neuromodulation of the NAc can enhance cognition in psychiatric disorders^25,26^. Building on this evidence, a previous TIS study targeting the NAc in patients with bipolar disorder also reported cognitive improvement, further underscoring the therapeutic potential of NAc-focused neuromodulation in addressing cognitive deficits in these disorders^13^. Despite this consistency, an interesting divergence exists: whereas the bipolar disorder study utilized a 40 Hz beat frequency, our protocol employed 130 Hz stimulation, and both frequencies were associated with improved cognitive function in their respective patient populations. The potential reasons for this discrepancy remain unclear, but we hypothesize that it may reflect differences in how the NAc contributes to the pathophysiology of SCZ versus bipolar disorder. This observation suggests that disease-specific frequency parameters may be required to optimally engage distinct dysfunctional neural circuits.

The present study demonstrated that TIS exhibited a favorable safety and tolerability profile, with only mild and transient adverse effects. The most frequently reported adverse event was “electrode sinking”, “transient current sensations”, and fatigue, which is consistent with previously reported findings in human TIS studies^7^. It is noteworthy that some participants received stimulation intensities that exceeded those employed in conventional protocols. However, even at the highest current intensity of 4 mA, TIS was well tolerated, as reflected by a mean VAS score of 7.6 ± 1.0. These results further support the safety and feasibility of TIS as a non-invasive neuromodulatory approach.

In this study, several innovative features warrant discussion. Firstly, the findings demonstrate that TIS applied at currents exceeding general TIS intensities (2.0 - 2.5 mA) remains safe and well-tolerated. Previous computational and animal studies indicate that TIS produces weaker electric field intensities than tACS at identical stimulation amplitudes^27,28^. This suggests that higher current intensities may be required to achieve therapeutic field strengths, while the findings of this study provide empirical justification for the safety of such elevated TIS protocols. Secondly, this study employed individualised MRI-based electric field modelling, which enhances targeting precision by minimising spatial bias due to inter-individual differences in brain anatomy. Thirdly, video recordings were made of the initial intervention session and the process of determining stimulation intensity for each participant, thus providing an objective reference for the reproducibility of treatment and methodological transparency.

This study has several limitations. First, this study is a single-arm non-randomized trial. Although strict control of patients’ medication during the study minimized bias, randomized controlled trials are needed to verify our findings. Secondly, the relatively limited sample size may restrict the generalizability of the results, thus necessitating further research with larger sample size. Finally, the follow-up period in this study was limited to two weeks. Future studies should incorporate longer-term follow-up to elucidate the durability of TIS treatment effects in patients with SCZ.

In conclusion, our findings indicate that TIS is a feasible and safe novel noninvasive DBS technique for patients with SCZ. By targeting the NAc, TIS may alleviate the negative symptoms and cognitive dysfunction in SCZ patients.

## Data Availability

All data produced in the present study are available upon reasonable request to the authors

## Data availability

The datasets generated and analysed during the current study, including original treatment videos, are not publicly available to protect participant privacy. De-identified data may be made available upon reasonable request to the corresponding author, subject to a signed data access agreement and ethical approval.

## Acknowledgments

We thank Liwen Feng from NeuroDome Clinical Technology Co., Ltd. for his technical assistance in data processing during this research. This work is supported by the Natural Science Foundation of Tianjin (Grant No. 24JCYBJC01000). The Fig 1. is created in BioRender.com.

## Reference

1. Kahn RS, Sommer IE, Murray RM, et al. Schizophrenia. Nature reviews Disease primers. Nov 12 2015;1:15067. doi:10.1038/nrdp.2015.67

2. Wang Y, Zhang C, Zhang Y, et al. Habenula deep brain stimulation for intractable schizophrenia: a pilot study. Neurosurg Focus. Jul 2020;49(1):E9. doi:10.3171/2020.4.FOCUS20174

3. Francis MM, Hummer TA, Vohs JL, et al. Cognitive effects of bilateral high frequency repetitive transcranial magnetic stimulation in early phase psychosis: a pilot study. Brain Imaging Behav. Jun 2019;13(3):852–861. doi:10.1007/s11682-018-9902-4

4. Mikell CB, Sinha S, Sheth SA. Neurosurgery for schizophrenia: an update on pathophysiology and a novel therapeutic target. Journal of neurosurgery. Apr 2016;124(4):917–28. doi:10.3171/2015.4.Jns15120

5. Corripio I, Roldan A, Sarro S, et al. Deep brain stimulation in treatment resistant schizophrenia: A pilot randomized cross-over clinical trial. EBioMedicine. Jan 2020;51:102568. doi:10.1016/j.ebiom.2019.11.029

6. Fenoy AJ, Simpson RK, Jr. Risks of common complications in deep brain stimulation surgery: management and avoidance. Journal of neurosurgery. Jan 2014;120(1):132–9. doi:10.3171/2013.10.JNS131225

7. Yang C, Xu Y, Feng X, et al. Transcranial Temporal Interference Stimulation of the Right Globus Pallidus in Parkinson’s Disease. Mov Disord. Aug 12 2024;doi:10.1002/mds.29967

8. Grossman N, Bono D, Dedic N, et al. Noninvasive Deep Brain Stimulation via Temporally Interfering Electric Fields. Cell. Jun 1 2017;169(6):1029–1041 e16. doi:10.1016/j.cell.2017.05.024

9. Violante IR, Alania K, Cassara AM, et al. Non-invasive temporal interference electrical stimulation of the human hippocampus. Nat Neurosci. Nov 2023;26(11):1994–2004. doi:10.1038/s41593-023-01456-8

10. Wessel MJ, Beanato E, Popa T, et al. Noninvasive theta-burst stimulation of the human striatum enhances striatal activity and motor skill learning. Nat Neurosci. Nov 2023;26(11):2005–2016. doi:10.1038/s41593-023-01457-7

11. Zheng S, Fu T, Yan J, et al. Repetitive temporal interference stimulation improves jump performance but not the postural stability in young healthy males: a randomized controlled trial. J Neuroeng Rehabil. Mar 20 2024;21(1):38. doi:10.1186/s12984-024-01336-7

12. Ma R, Xia X, Zhang W, et al. High Gamma and Beta Temporal Interference Stimulation in the Human Motor Cortex Improves Motor Functions. Front Neurosci. 2021;15:800436. doi:10.3389/fnins.2021.800436

13. Zhou H, Wang M, Qi S, et al. Efficacy and Safety of Transcranial Temporal Interference Stimulation for Treating Bipolar Disorder with Depressive Episodes. medRxiv. 2024:2024.11.19.24317540. doi:10.1101/2024.11.19.24317540

14. Alania K, Borella J, Violante I, et al. Non-Invasive Temporal Interference Hippocampal Stimulation in Early Alzheimer’s Disease. Alzheimer’s & Dementia. 2023;19(S21):e077433. 10.1002/alz.077433

15. McCutcheon R, Beck K, Jauhar S, Howes OD. Defining the Locus of Dopaminergic Dysfunction in Schizophrenia: A Meta-analysis and Test of the Mesolimbic Hypothesis. Schizophrenia bulletin. Oct 17 2018;44(6):1301–1311. doi:10.1093/schbul/sbx180

16. Kapur S. Psychosis as a state of aberrant salience: a framework linking biology, phenomenology, and pharmacology in schizophrenia. The American journal of psychiatry. Jan 2003;160(1):13–23. doi:10.1176/appi.ajp.160.1.13

17. Gault JM, Davis R, Cascella NG, et al. Approaches to neuromodulation for schizophrenia. J Neurol Neurosurg Psychiatry. Jul 2018;89(7):777–787. doi:10.1136/jnnp-2017-316946

18. Seamans JK, Yang CR. The principal features and mechanisms of dopamine modulation in the prefrontal cortex. Prog Neurobiol. Sep 2004;74(1):1–58. doi:10.1016/j.pneurobio.2004.05.006

19. Kay SR, Fiszbein A, Opler LA. The positive and negative syndrome scale (PANSS) for schizophrenia. Schizophrenia bulletin. 1987;13(2):261–76. doi:10.1093/schbul/13.2.261

20. Fabri A, Giezeman G-J, Kettner L, Schirra S, Schönherr S. On the design of CGAL a computational geometry algorithms library. Software: Practice and Experience.2000;30(11):1167–1202.10.1002/1097-024X(200009)3 0:11<1167::AID-SPE337>3.0.CO;2-B

21. Dular P, Geuzaine C, Henrotte F, Legros W. A general environment for the treatment of discrete problems and its application to the finite element method. IEEE transactions on magnetics. 1998;34(5):3395–3398.

22. Rolls ET, Huang CC, Lin CP, Feng J, Joliot M. Automated anatomical labelling atlas 3. NeuroImage. Feb 1 2020;206:116189. doi:10.1016/j.neuroimage.2019.116189

23. Andreasen NC. The Scale for the Assessment of Negative Symptoms (SANS): conceptual and theoretical foundations. Br J Psychiatry Suppl. Nov 1989;(7):49–58.

24. Nuechterlein KH, Green MF, Kern RS, et al. The MATRICS Consensus Cognitive Battery, part 1: test selection, reliability, and validity. The American journal of psychiatry. Feb 2008;165(2):203–13. doi:10.1176/appi.ajp.2007.07010042

25. Floresco SB. The nucleus accumbens: an interface between cognition, emotion, and action. Annual review of psychology. Jan 3 2015;66:25–52. doi:10.1146/annurev-psych-010213-115159

26. Cauda F, Cavanna AE, D’Agata F, Sacco K, Duca S, Geminiani GC. Functional connectivity and coactivation of the nucleus accumbens: a combined functional connectivity and structure-based meta-analysis. Journal of cognitive neuroscience. Oct 2011;23(10):2864–77. doi:10.1162/jocn.2011.21624

27. Vieira PG, Krause MR, Pack CC. Temporal interference stimulation disrupts spike timing in the primate brain. Nature communications. May 29 2024;15(1):4558. doi:10.1038/s41467-024-48962-2

28. Karimi N, Amirfattahi R, Zeidaabadi Nezhad A. Neuromodulation effect of temporal interference stimulation based on network computational model. Front Hum Neurosci. 2024;18:1436205. doi:10.3389/fnhum.2024.1436205

