## Supplementary figures and images for "Efficacy and Safety of Transcranial Temporal Interference Stimulation for Improving Negative Symptoms and Cognition in Schizophrenia: A Pilot Study"

### Fig.S1.jpg

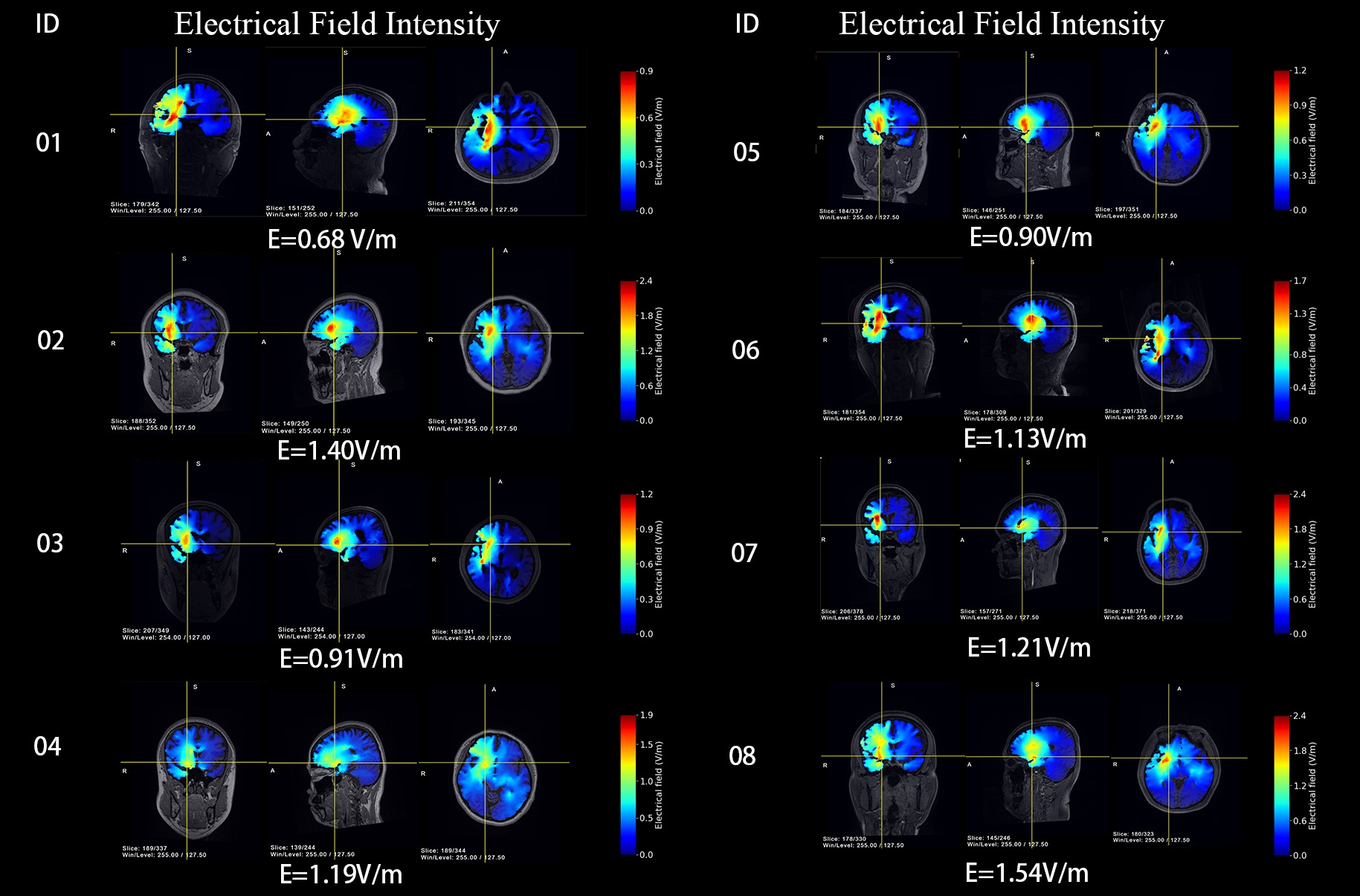

### fig.S2.tif

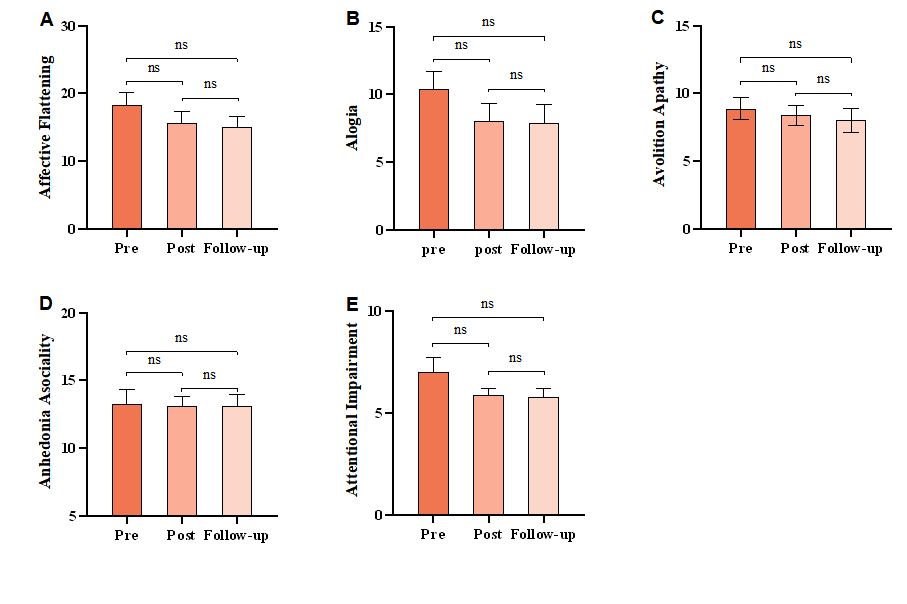
