## Supplemental Table 1 for "Efficacy and Safety of Transcranial Temporal Interference Stimulation for Improving Negative Symptoms and Cognition in Schizophrenia: A Pilot Study": Table S1.docx

| Patient | Electrodes Placement | |  | Current Amplitude (mA) | |  | Amplitude Ratio (I1: I2) |
| --- | --- | --- | --- | --- | --- | --- | --- |
|  | I1 | I2 |  | I1 | I2 |  |  |
| 1 | AF4(+), Fz(-) | FT10(+), O10(-) |  | 2.50 | 2.70 |  | 1.85 : 2.00 |
| 2 | F8(+), F9(-) | T8(+), O10(-) |  | 3.00 | 3.75 |  | 1.60 : 2.00 |
| 3 | F6(+), C1(-) | F10(+), O9(-) |  | 2.30 | 3.50 |  | 1.30 : 2.00 |
| 4 | AF8(+), FP1(-) | FT8(+), O9(-) |  | 3.00 | 2.60 |  | 2.00 : 1.50 |
| 5 | AF8(+), FP1(-) | T8(+), O9(-) |  | 3.00 | 3.00 |  | 1.00 : 1.00 |
| 6 | F8(+), P6(-) | F10(+), FT7(-) |  | 4.00 | 3.40 |  | 2.00 : 1.70 |
| 7 | F4(+), OZ(-) | F8(+), O10(-) |  | 4.00 | 2.80 |  | 2.00 : 1.40 |
| 8 | FPz(+), T7(-) | FT8(+), P8(-) |  | 4.00 | 4.00 |  | 1.00 : 1.00 |
| I1: primary pair electrode; I2: secondary pair electrode. Electrodes placement are determined by the 10-10 EEG system. | | | | | | | |

Table S1, Stimulation parameters and electrode configurations
